# Preservation of cognition in hypertension-treated South Indian rural population

**DOI:** 10.1101/2020.01.28.20019125

**Authors:** Radhika Rajkumar, Alex Divya Merciline, Suresh Kumar Muthukrishnan, Murali Subhashree, Muniswamy Duraimurugan, Velmurugan JanakiDevi, Manjula Datta, Jamuna R. Subramaniam

## Abstract

Change in diet, life style and increased life expectancy has led to the dramatic escalation in old age related complication like cognitive decline leading to dementia. Cardiovascular diseases (CVD) are huge risk factors for dementia, including Alzheimer disease (AD). Hypertension is very well known to cause cognitive impairment. Control of CVD could provide protection against dementia. Earlier, in the mouse model of AD, reserpine, an antihypertensive and antipsychotic drug could elicit improvement in the working memory in AD model mice and enhance the same in normal mice. Hence, Cognitive protection in the patients on chronic antihypertensive drug which contains reserpine was evaluated. Cognition in a cohort (in the South Indian rural population) of hypertensive patients (majority age group – 50-70 years) who have been chronically treated with a combinatorial drug, (adelphane/adelphane esidrex-Novaritis, Switzerland) consisting of reserpine and hydrazine for years was compared with controls without hypertension. The cohorts were age, sex, socio-economic, education background matched. Cognition was scored using the Tamil version of: Addenbrooke’s Cognitive Examination-III (T-ACEIII) and Tamil-Montreal Cognitive Assessment (T-MoCA) scales. The composite T-ACEIII score of control and treated groups were 53.6 and 53.2, respectively. T-MoCA scores (Control-15.1 and Treated-14.7) did not show much alteration. Further, the mean scores of the control and treated groups’ individual components of cognition in ACE, namely, Attention, Memory, Fluency, Language and visuospatial – cognitive skills also did not reveal significant difference. Thus, controlling blood pressure or hypertension with chronic antihypertensive medication like adelphane/adelphane esirdex (reserpine containing drugs) has retained normal cognition in both genders.

## Introduction

The world is aging at an alarming rate^1^. The increase in life expectancy has led to a world enriched with greying population which will increase from 12% to 22% in the next two-three decades^2^. In 2050, 90% of the people above 60 years will be living in the developing countries. Non –communicable chronic diseases like diabetes^2^, hypertension^3^ and neurodegenerative diseases are directly proportional to aging^4^. Cognition declines with age and the neurodegenerative disease, Alzheimer disease (AD)-loss of memory (dementia) has become a huge geriatric burden with 1 in 10 above 65 years developing the disease. Further, two thirds of the AD patients are women^5^. There is hardly any treatment for AD. Acetylcholine inhibitors, donepil^6,7^ and glutamate’s NMDA receptor partial antagonist, memantine^6,8^ are the only drugs available. They are of very limited use and given after the onset of the disease and mostly palliative in nature. The molecular changes that lead to AD development starts more than a decade ahead of the disease manifestation^9^. Hence, the etiology needs to be identified and preventive strategy be initiated from mid–life to delay/protect against cognitive decline.

Some of the risk factors for AD are cardiovascular diseases^10^ and diabetes. Of these, hypertension seems to contribute to cognitive decline significantly. More importantly, one third of the world population in general and 66% above age 65 suffer from hypertension^11,12^. One of the major reasons for cognitive decline is impairment of blood circulation to the brain, which is known to cause both vascular dementia and Alzheimer disease^13,14^. The prevalence of hypertension in low and middle income countries including India is 40% ^15^. Therefore diagnosis of hypertension and its treatment is essential. Drugs targeting different pathways^16^, namely, calcium channel blockade(nitrendipine)^16,17^, angiotensin-renin system’s angiotensin converting enzyme inhibitors-enalapril, and or angiotensin receptor II (ARII) inhibition, with or without a diuretic (hydrazine/ hydrochlorothiazide), could control hypertension^15^. The longer the time on antihypertensive, lesser is the chance of dementia^18^. Interestingly, various large cohort meta-analysis has revealed, that calcium channel blockers and ARII inhibitors may protect better against cognitive decline^16^. Another trial which included the biogenic amine downregulator, reserpine, was inconclusive due to several control group switching to treatment^16^.

Independently, in a *C. elegans* screen, reserpine was found to extend lifespan^19^, albeit, through modulation of acetylcholine release^20^, partially through the action of Goi coupled dopamine receptor, *dop-3* ^21^. More importantly, reserpine could protect against the Alzheimer disease causing Amyloid beta toxicity induced paralysis in *C. elegans*^22^ and improved working memory in the 5XFAD Alzheimer disease model mice^23,24^ suggesting reserpine as a potential drug to provide cognitive protection.

Starting from 1995, Kidney Help Trust, a Community Health initiative, had screened the entire population of ∼56 villages and hamlets in an around Chennai, a rural South Indian population in the state of Tamil Nadu, India for both diabetes and hypertension^25-29^. Hypertension was treated with Adelphane which consists of reserpine (0.1mg) and a diuretic -hydralazine (10mg) ^25-30^ or Adelphane esirdex ^25-29^. As hypertension is relentless and needs long-term medication, these patients had to remain on reserpine for several years.

Reserpine ^25-29^ was the antihypertensive^31,32^ drug of choice in this community health initiative and for the Chinese rural population^33^. Reserpine is a historical entity, of plant origin, specifically, an alkaloid purified from the roots of *Raulwoflia serpentina* (common name -snake root; colloquial name -sarpagandha). It was used in Ancient ayurvedic medicine for the treatment of hypertension, insomnia, as tranquilizer, snake bites and insanity^34-36^. Later, reserpine was shown to provide protection in hypertension^31,37^ and psychosis along with chlorpromazine^32,38, 39^.

Currently, reserpine is an FDA approved drug for second line treatment of hypertension and treatment resistant psychosis, though availability is a challenge. In rural China, more than 44% use antihypertensive herbal compounds preparation containing reserpine^33^. Reserpine acts through downregulation of biogenic amines (dopamine, serotonin, epinephrine and norephineprine) neurotransmitters at the synapse. It inhibits these biogenic amines loading into synaptic vesicles by irreversible inhibition of vesicular monoamine transporter (VMAT) ^40-42^. Reserpine was a very effective and inexpensive drug for a huge population in low and middle income countries.

Identification of whether control of hypertension with reserpine or adelphane/adelphane hydrilex will provide the added benefit of cognitive protection. This will be extremely beneficial to the aging economically weaker community to have a dignified life and protect the society against huge social and financial liability. Given the importance, we address the cognitive benefits of hypertension treatment with reserpine as a combinatorial drug with a diuretic (due to non-availability reserpine as a single drug) in the South Indian context, as we found reserpine’s ability to enhance cognition in the 5XFAD Alzheimer disease mouse model and normal mice^22^.

## Materials and Methods

### Study Design

A retrospective cross sectional observational study was carried out about the cognitive abilities of the population with hypertension and treatment in comparison with matched normal volunteers.

### Subjects

Age, sex, education and socioeconomic background matched control (not hypertensive) and hypertensive patients on Adelphane/ Adelphane Esidrex from Novaritis, Basel, Switzerland for several years were recruited. The International Standard Sri Ramachandra University Institute Ethics Committee clearance was obtained before the study was initiated. Further, patients informed consent was acquired. Both the group cohorts were from the same locality. The control group (N= 107) and the Adelphane (reserpine) treated group (N=106). Most of the population had basic education of being able to read and write tamil at the basal level at least words. The highest education was passing elementary or high school which was very few. This was a blinded cross-sectional study. Further, all methods were carried in accordance with the relevant guidelines.

### Period

The field visits and data collection took one year. Further, data consolidation and analysis took another six months. On the whole, it was started in May 2015 and ended in December 2016.

### Hypertension treatment

The treatment details are already reported in Rajkumar *et al*., (2018)^43^. Briefly, The <u>Kidney Help Trust of Chennai</u> provided reserpine in the form of Adelphane (0.1 mg reserpine and 10 mg of hydralazine)/Adelphane Esidrex (has 10 mg of hydrochlorothiazide added to Adelphane) of Novaritis, Switzerland. The hypertension patients, South Indian population in 56 villages and hamlets around Chennai, Tamil Nadu, India were treated as detailed above from 1995 onwards. The dosage varied from 1 tablet to 4 tablets for a day according to the patient status. Management of hypertension with this drug was very effective. Fixed specific dose worked well for a very long period (in years) in the patients. In addition, it was well tolerated.

### Cognition evaluation

#### Addenbrooke’s cognitive Examination (ACE-III) screening

Permission was obtained to use ACE-III before the scale was tested on the cohorts. The worldwide well accepted Addenbrooke’s cognitive examination revised version –III^44, 45^ was adapted to the Indian local language – Tamil (one of the Ancient living language) of the state Tamil Nadu and administered to the control and patients in a blinded manner. The individual cognitive domains and the total ACE-III-T scores were obtained.

### MoCA Screening

The freely available Montreal cognitive assessment scale (MoCA)^45,46^ developed in Canada in 1996 translated in the native local language, Tamil, gauges the individual cognitive domains like language, visuospatial abilities, memory and recall and abstract thinking. This gives a consolidated total score (Maximum= 30) for control and the patients cohorts. This is referred to as MoCA-T.

### Statistical analysis

Statistical analyses of the scores were done using SigmaPlot 10.0. Statistical significance was determined by Students t-test and Mann-Whitney Rank Sum Test.

## Results

Control group (107) and patients receiving adelphane/adelphane esirdex (106) were objectively screened for Cognitive abilities using the universally accepted and adapted ACE-III^45^ and MoCA^45^ scales. More importantly, these scales were adapted to the local language, Tamil, the mother tongue of all the participants. The results obtained were analyzed for individual cognitive domains and the consolidated total scores.

### Cognitive protection by antihypertensive drug adelphane/adelphane esirdex

The consolidated ACE-III-T (Fig. 1; Table 1) and MoCA-T (Fig.1B) scores of control and treated groups independent of age and gender did not show significant difference. The mean values foe ACE-III-T for control and treated are 53.6 and 53.2, respectively, for a total score of 100. For MoCA-T, the mean consolidated scores are: Control-15.1 and Treated-14.7 out of a total score of 30. Lack of significant difference between the control and treated group suggests that anti-hypertensive treatment has retained the cognition in the hypertension patients on adelphane/adelphane esirdex.

**Table -1.**
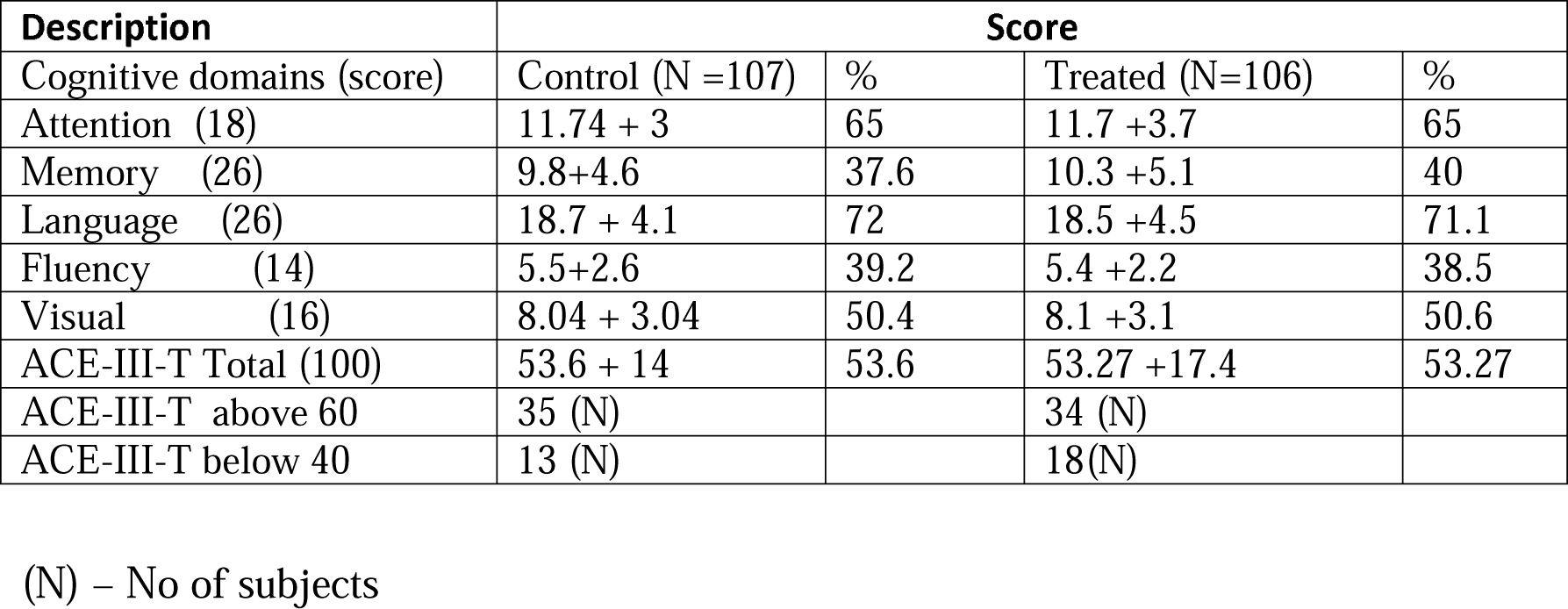
Addenbrooke’s (ACE-III-T) Cognitive Score.

**Fig. 1.**
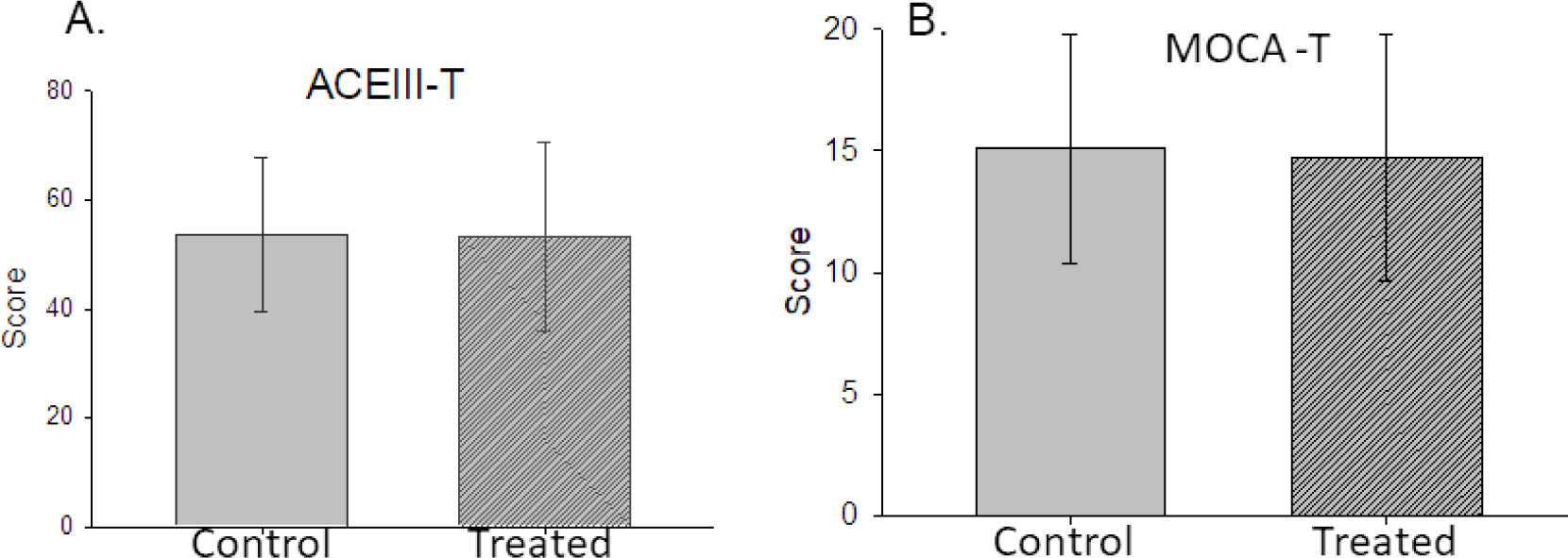
Chronic Antihypertensive treatment maintains cognition. A. The total ACEIII-T score of normal (control) and patients on antihypertensive (PAH). B. The total MOCA-T score of the normal and PAH.

### Protection in different cognitive domains

Next, we determined the mean scores in the individual cognitive domains, namely, attention, memory, fluency, visual-spatial and language obtained with ACE-III-T. We find no significant difference between the control and treated group (Fig. 2; Table 2). Antihypertensive treatment is able to protect individual cognitive domains as well.

**Fig. 2.**
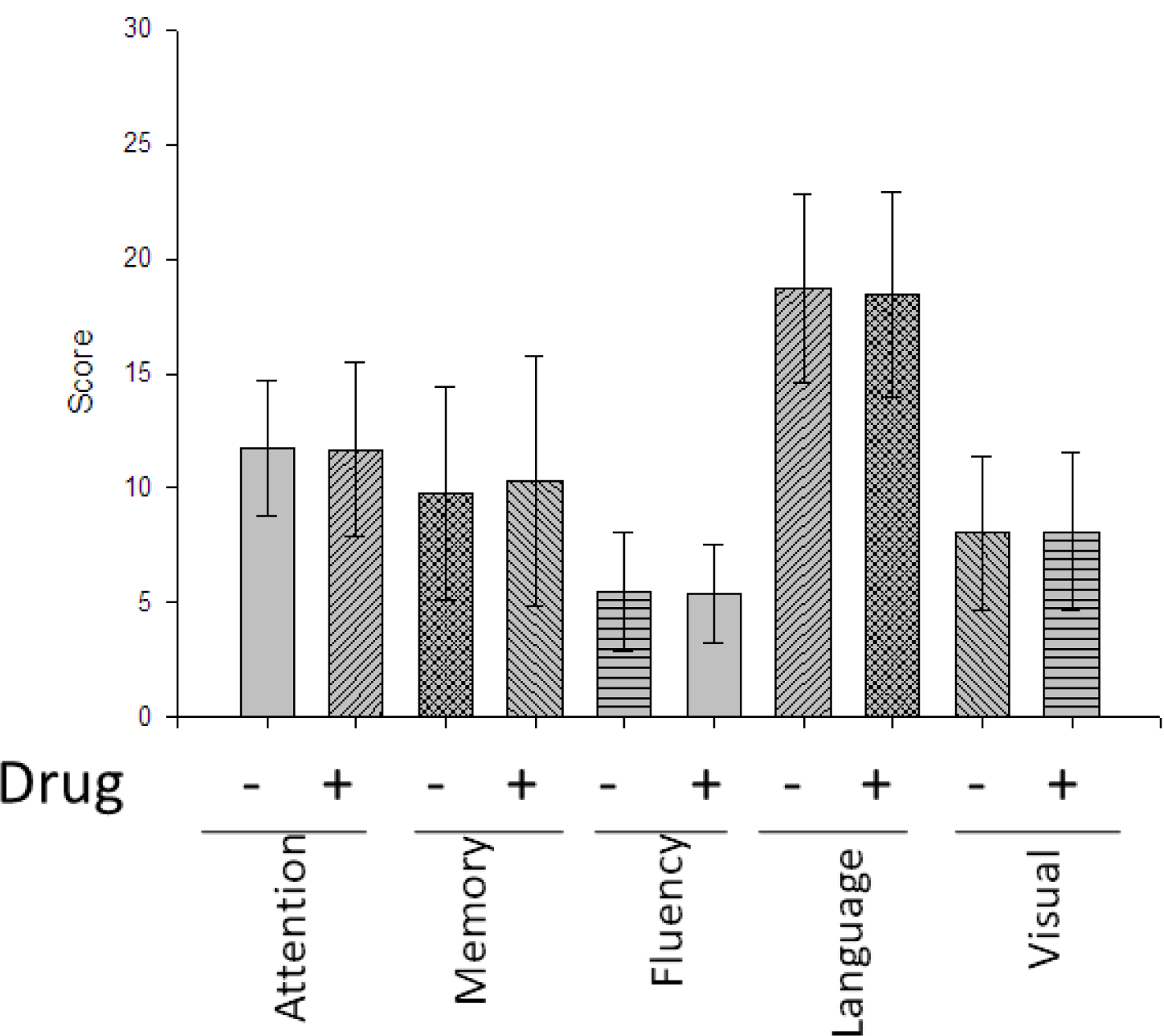
Chronic antihypertensive regimen retains normalcy in various cognitive domains. The cognitive domains measured in ACE-III-T as (x-axis) Attention, Memory, fluency, language and visuo-spatial skills scores (Y-axis) remains normal in PAH.

**Table:** Agewise comparison of cognitive domains with and without antihypertensive drug regimen.

| Age group<br>in years | Attention |  | Memory |  | Fluency |  | Language |  | Viso-spatial |  | ACE- Total |  |
| --- | --- | --- | --- | --- | --- | --- | --- | --- | --- | --- | --- | --- |
|  | Control | Treated | Control | Treated | Control | Treated | Control | Treated | Control | Treated | Control | Treated |
| 30-50 | 12.5 ± 2.8 | 12.1±3 | 11 ±5.1 | 12.9±5.8 | 6.5 ±2.9 | 5.9 ± 2.4 | 20.2 ±3.8 | 20.8±3.6 | 9 ±3.3 | 9.6±3.7 | 57.2±14.1 (19) | 61.2±14.4 (14) |
| 51-60 | 11.7 ± 3 | 12.8±2.9 | 11.1 ± 4.6 | 11.3±4.9 | 5.8 ± 3 | 5.6 ± 2.3 | 20 ±2.5 | 20 ±3.5 | 9.2 ± 3.5 | 8.8 ±3.3 | 57.2±12.8 (27) | 58.6 ±13.1 (31) |
| 61-70 | 12 ±3.1 | 10.8±4.2 | 9.3 ±4.8 | 9.3±5.1 | 5.1±2.4 | 5.2 ±1.8 | 18.8±3.9 | 16.8±4.7 | 7.5 ± 3.2 | 7.2 ±3.4 | 52.6±14.3 (39) | 49.4±16.2 (37) |
| 71-80 | 10.7 ±3 | 11.4±3.9 | 7.5±3.5 | 9.2±5.8 | 4.9±2.1 | 5.4±2.5 | 15.8±4.6 | 17.7±4.8 | 6.9±3.2 | 7.5±3.2 | 45.5±11.2 (19) | 51.4±17.5 (22) |
| 81-90 | 12±2.6 | 10.5±9.2 | 9.3±3.5 | 6.5±9.2 | 4.7±2.5 | 5±1.4 | 15±7.2 | 16±4.2 | 7±4 | 9.5±4.9 | 48±18.5 (3) | 47.5±29 (2) |

ACE-III-Tamil -total and the individual domains, attention, memory, fluency and visuo-spatial comparison based on age with(Treatment) antihypertensive treatment and without(control) are given. ± Standard deviation (S.D) is given.

### Cognitive protection across various ages

Further, we analyzed the individual cognitive domains scores segregated based on age (Fig. 3&4; Table 2). Though there is a marginal decrease in cognitive scores with age, adelphane/adelphane esirdex treatment is able to maintain it at the same levels as the controls when compared within the specific age group.

**Fig. 3.**
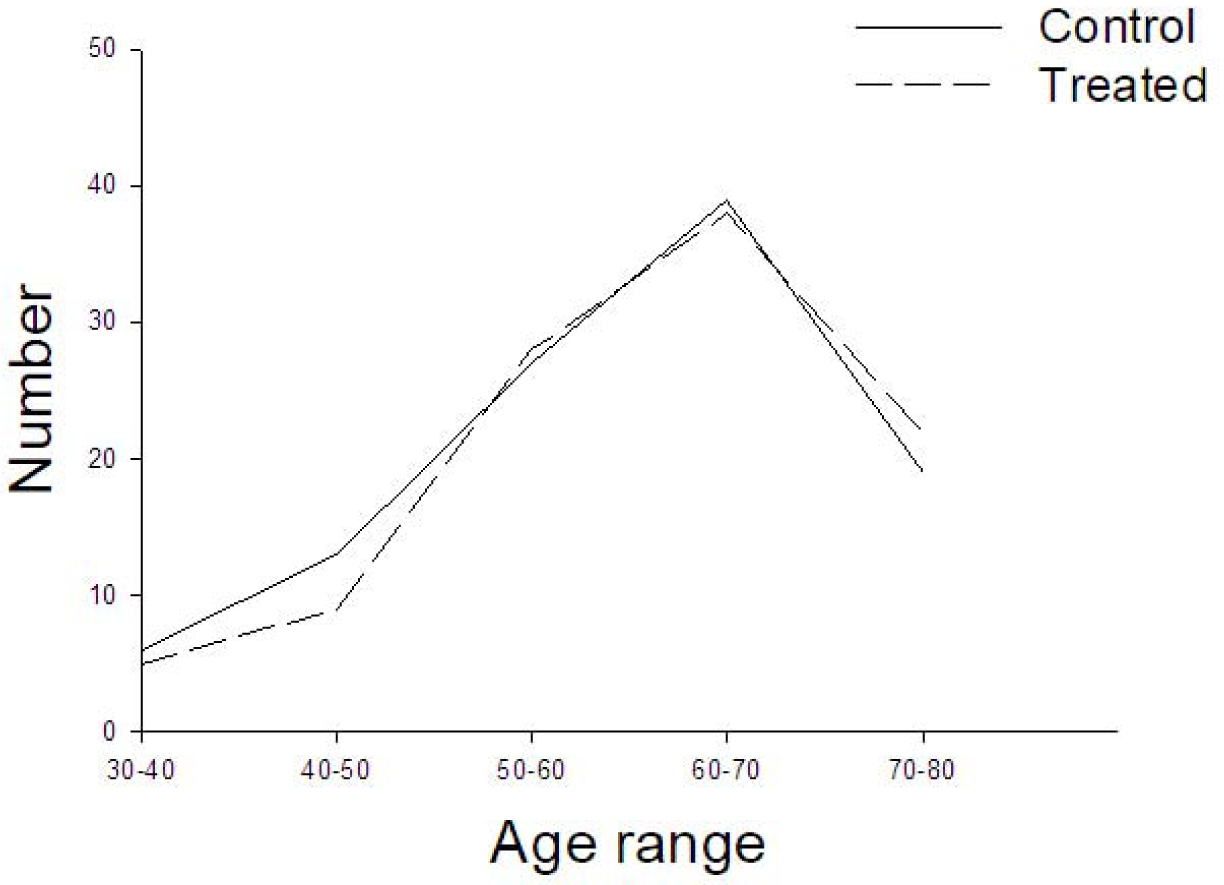
Age distribution of the study cohorts. Control – Black line: PAH-line with dashes

### Cognition in males and females

When the cognitive ability protection by antihypertensive treatment was measured in both the genders, reserpine works effectively in both males and females (Fig.5; Table 3). Here again, a significant difference was not noticed for either gender compared to the control.

**Table -3.** Gender wise comparison of cognitive domains of ACE-III-T.

| Gender | Attention |  | Memory |  | Fluency |  | Language |  | Visuo-spatial |  | ACE -Total |  |
| --- | --- | --- | --- | --- | --- | --- | --- | --- | --- | --- | --- | --- |
|  | Control | Treated | Control | Treated | Control | Treated | Control | Treated | Control | Treated | Control | Treated |
| **Female** | 11.2±2.7 | 10.8±3.3 | 9.3±4.6 | 9.5±4.9 | 5.2±2.6 | 5.1±1.8 | 17.9±4.0 | 17.5±4.3 | 7.3±3.2 | 7.3±3.2 | 50.8 ± 13.3<br>(n=80)* | 50.5 ± 14.3<br>(n=78)* |
| **Male** | 13.1±3.2 | 13.9±3.8 | 11.1±4.5 | 12.5±6.0 | 6.4±2.1 | 6.2±2.5 | 21.1±3.3 | 20.8±3.8 | 10.0±2.8 | 10.2 ± 3.1 | 61.8 ± 12.7<br>(n=27)* | 63.7±16.6<br>(n=28)* |

\*n– number of subjects is the same for ACE total and the individual domains.

### Cognitive comparison between males and females

An independent observation shows slightly less mean cognitive score as per ACE-III-T among women (51%) than men (64%)(Table 3). Further, strangely, there was a significant difference in cognition between genders with males mildly better than females in the Language, Fluency and visuo-spatial skills and overall ACEIII-T (Fig. 5; Table 3) (13% better).

**Fig. 4.**
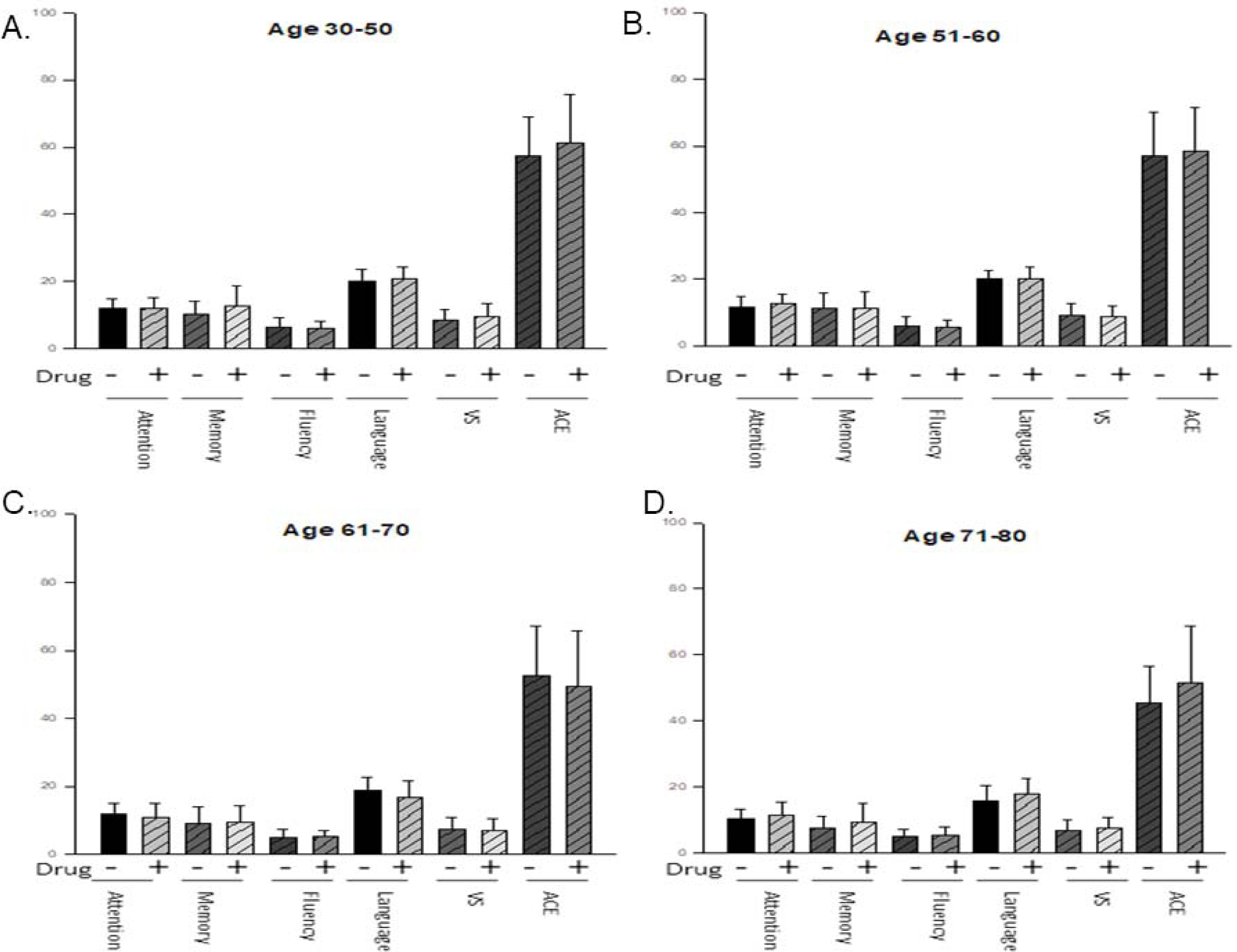
Different cognitive domains are normal across various ages upon antihypertensive treatment. Comparison between normal and PAH of different age groups A. 30-50; B. 51-60; C. 61-70; D. 71-80. Y-axis –scores; X-axis – control and treated – specific cognitive domains and total ACE-III-T.

**Fig. 5.**
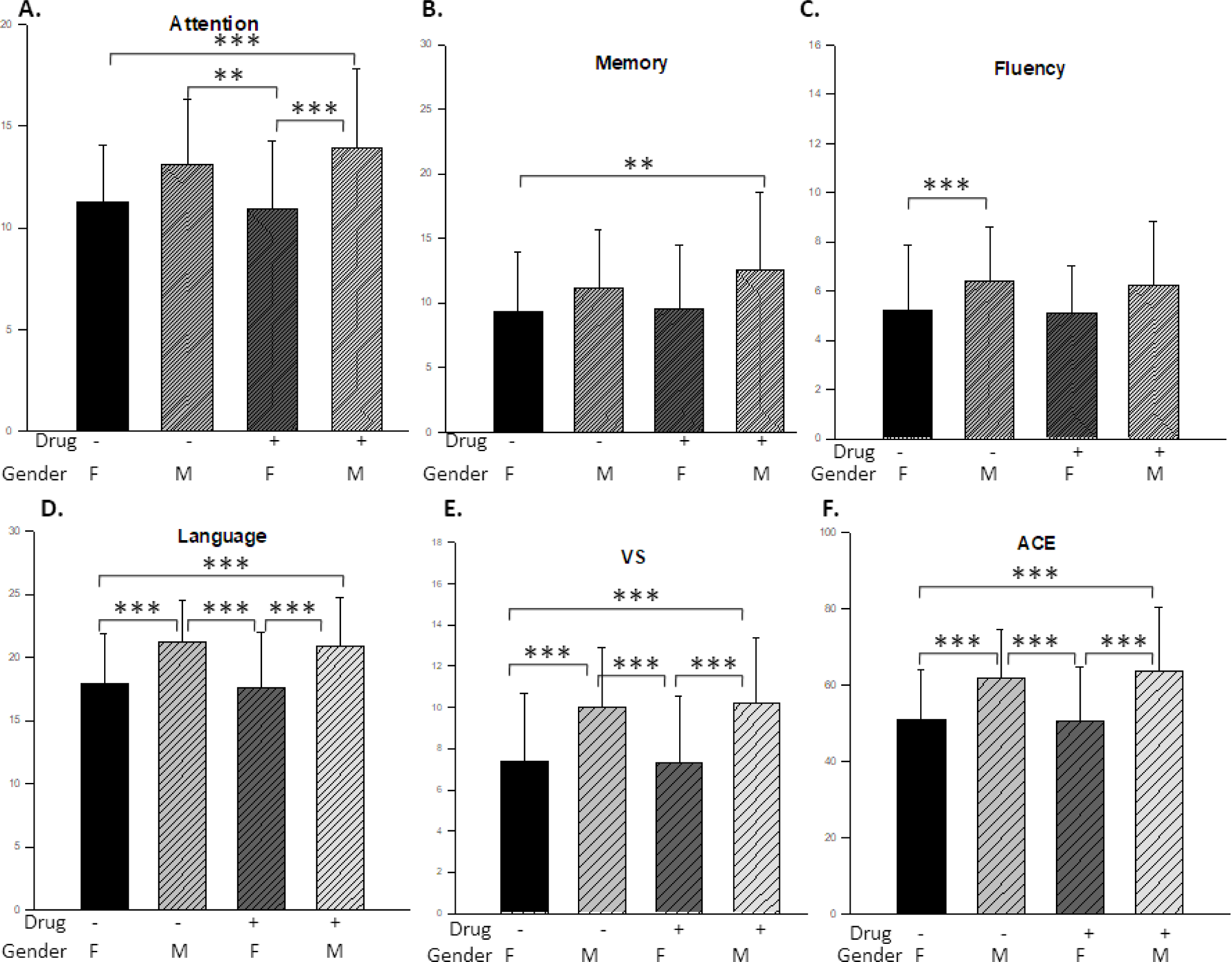
Chronic antihypertensive treatment protects in both genders. Comparison of the cognitive domains between normal males and females with the respective PAH. Y-axis – Scores: X axis-Drug treatment and gender. A. Attention; B. Memory; C. Fluency; D. Language; E. Visuo-spatial skill; F. Total score in ACE-III-T.

Thus, the reserpine containing combinatorial antihypertensive drug, Adelphane/adelphane esidrex was able to protect cognition across different cognitive domains. Hence, antihypertensive drug treatment is one modality for preservation of cognition in the geriatric population when started from middle age onwards.

## Discussion

Hypertension is one the most prevalent (30-40%) and silent killer, non-communicable diseases which can occur at any age, more so in middle age ^16, 47^. More importantly, middle age hypertension shows very strong association with later age dementia^16,48^. Hence, hypertension alone and in combination with diabetes and dementia with huge morbidity and mortality rates, and the social, financial and economic burden on the world community, needs to be contained in all its totality. Towards this, Kidney Help Trust in Chennai, Tamil Nadu, India in 1995 started a major initiative to screen the rural population for diabetes and hypertension and providing treatment for persons with these diseases^25-29^. The antihypertensive drug used is Adelphane/adelphane esidrex. This was the least expensive hypertension medication and also the most efficient, effective at the same dosage for years and well tolerated, given that antihypertensive drugs need to be taken lifelong and cost was a major factor. Hence, reserpine was the drug of choice for the poor ^25-29^. Sadly, midway through the study due to non-availability of adelphane/adelphane esidrex, the patients were switched to arkamin.

As there is a correlation between hypertension and cognitive decline, the cognition of the hypertension patients on the antihypertensive drug adelphane/adelphane esidrex in comparison with normal age, sex and socioeconomic background matched subjects was evaluated. The mean blood pressure was reduced from 149/88 to 143/83 upon chronic antihypertensive treatment^43^. Screening of various cognitive domains like memory, language, fluency, attention and visuospatial skills using the worldwide accepted ACE-III scale translated in the local language Tamil (ACE-III-T) revealed preservation of cognition across all the domains in all the age groups in both genders (Fig. 1-5, Tables 1-3). The ACE-III-T mean total score was 53.6(control) and 53.2(treated) for 100. Similarly, no difference was noticed in the screening with MOCA-T 15.1 control and 14.7 (treated) for 30. Cognition was fully preserved in all the age groups in both the genders when compared with the control (Fig. 3 & 4, Fig. 5; Table 2 & 3).

One of the major challenge was administration of the cognitive scales in the rural population where most of them had only basic education^49^. To the best of our knowledge, such cognitive screening has not been carried out in the South Indian rural population. Further, there seems to be a correlation between education and the cognitive score when such a study was carried out using ACE-III translated in another Indian language, Malayalam^50^. In addition to the total scores, the individual domains were also screened. In this again, no difference was noticed between control and treated groups in both genders. But, further analysis of ACE-III-T scores revealed information like, i) with age cognition slightly declines (Fig. 4 A-D, Table 2) in all the domains. ii) Males have slightly higher ACE-III-T score than females (Fig.5; Table 3). One of the reason could be lack of intellectual pursuits in the women of the age group that was tested. As told by one of the woman participants (for the cognitive domains), she was not pursuing such activities after starting a family. In addition, as Alzheimer disease is more common in women, probably, more cognitive stimulation is needed. This needs to be seriously addressed.

Contrary to the negative publicity of reserpine causing suicidal tendency, reserpine is shown to work well as an antihypertensive drug in patients^32^and clear data for this negative effect also is lacking ^51^. Earlier for psychosis the dosage was in mgs while here only 0.1 mg to 0.4 mg is given. In the same population, depression screening by us has shown no increased incidence upon taking adelphane/adelphane esirdex^43^. Thus it is a safe drug, but now discontinued by Novaritis. Since Novaritis was supplying it for a long time, they may have their own reasons for discontinuing.

As it was a blinded study, 50% of control and adelphane/adelphane esirdex treated hypertensive patients were retraced. Of this, 80% of the patients were on this drug for more than two years and 33% for more than 10 years. A small population of 1.4% was on reserpine for more than 18 years. The blood pressure of control subjects was normal while the Adelphane/Adelphane esidrex treated patients it had been brought down by ∼10mm Hg in both diastolic and systolic blood pressure^43^. In contrast to the implicated negative effects, reserpine had been safe for chronic usage as an antihypertensive. To iterate this, 44% of rural Chinese population use reserpine containing herbal compound preparation^33^ for hypertension. In addition, reserpine had been an effective antihypertensive in older patients^52^.

In the current scenario of the world aging at a rapid rate, and the increase in mean lifespan, cognitive decline and Alzheimer disease in the geriatric population, reserpine could preserve cognition in the poor patients of age 50 and 70 of rural South Indian setting (Fig. 1 and Fig 2). The novelty of this study is the drug reserpine identified through the C*aenorhabditis elegans* lifespan extending^19^ and Alzheimer disease(AD) causing amyloid beta toxicity^22^ protecting effect, could provide cognitive enhancement in normal and 5XFAD transgenic AD model mice^23^ and reduce Aβ aggregates in another AD model mice Tg2576^24^ can preserve cognition in hypertensive patients (Fig.1). Given that reserpine is of ancient Ayurvedic medicine origin, the wisdom of the ancestors is praiseworthy.

## Data Availability

All the data has been consolidated and presented. This data was collected more than 3 years back. So, they may not be available.

## Acknowledgement

The authors are grateful to Dr. M.K. Mani, Managing Trustee, Kidney Help Trust Chennai for the encouragement, support and guidance and the employees of Kidney Help Trust who aided in the subjects recruitment and identification. The authors thank Prof. Sakthisekaran, Department of Community Medicine, Sri Ramachandra Institute of Higher Education and Research, for the discussions and suggestions. The authors have no conflict of interest.

## Note

As =Adelphane/Adelpane esidrex was not available, the hypertensive patients were switched to Arkamin midway through the study.

## Author Contribution

Conceptualization JRS; Methodology: SKM, ADM, MaD* and JRS; Validation-ADM; Consolidation – RR, ADM and MS; Formal analysis-MS, MD, VJD and JRS; Resources -JRS.; writing – original draft -JRS, review – all authors; editing -JRS; Administration and co-ordination – RR; Supervision-JRS and MaD*; MaD*-Manjula Datta

